# Comparing Vaccination Strategies in Canada Under Different Assumptions

**DOI:** 10.1101/2021.03.02.21252761

**Authors:** Timothy Ruse

**Affiliations:** Concordia University

**Keywords:** SARS-CoV-2, Vaccination, Canada, Pfizer

## Abstract

This paper estimates the outcomes of two different COVID-19 vaccination strategies in Canada for the mRNA vaccines currently approved for Emergency Use Authorization (EUA), modelled on the vaccination and effectiveness of the Pfizer vaccine which is likely to be more widely administered in Canada. The first strategy is the manufacturer recommended standard of two doses (two-dose strategy) given within 21 days apart versus a strategy of giving a larger group a single dose of vaccine (first-dose-for-most strategy) by delaying the second injection.

Three parameters are varied in the course of 36 estimation scenarios of the population-level effects of the two vaccination strategies. The first is the effectiveness of a single dose of vaccine at preventing disease, the second is the effectiveness of the vaccine at preventing transmission of the virus, and the third is the rate of transmission of the virus during the course of the simulations.

Over the course of the different scenarios, the first-dose-for-most strategy was superior in reducing disease transmission in all scenarios where vaccination is assumed to have an effect on viral transmission. The results for fatalities was mixed, with the first-dose-for-most strategy being superior in cases where a higher first-dose effectiveness at preventing disease was assumed.

Finally, in the best-guess scenarios where a 75% reduction in disease transmission and a 92.6% effectiveness at preventing disease from a single dose were used, the first-dose-for-most strategy was superior in a situation with reduced vaccine doses available, and switching to the first-dose-for-most strategy earlier helped to prevent a higher proportion of cases and deaths.

## MOTIVATION

The intent is to compare the effectiveness of the two strategies considered, given recent estimations of the effectiveness of a single dose of vaccine, and to explore the interactions of various parameters with respect to the effectiveness of those strategies. In the scenarios reported in this paper, three factors are varied:

1. The effectiveness of a single dose of vaccine at preventing disease (3 values)
2. The effectiveness of the vaccine (one dose or two) at preventing disease transmission (4 values)
3. The average number of people a person goes on to infect if no one were vaccinated (3 values)

Two timing related scenarios are also tested: one representing starting at an earlier point in time, and one where available vaccine doses are cut in half.

## PERIOD OF STUDY

The various scenarios are meant to simulate a 24 week period from the beginning of April, based on the assumption that any change in vaccination strategy would take some time to study and then implement. The number of vaccinations available during this period is taken from guidance provided by the Public Health Agency of Canada (PHAC), with the weekly distribution of vaccines estimated based on an increasing supply during the first 12 weeks followed by a steady distribution in the second 12 week period.

### CAVEATS

In the simulations, susceptibility to COVID-19 is entirely dependent on age, and the likelihood of transmitting of the virus onward is the same for everyone. The age-based susceptibility can be interpreted as a proxy for other risk-factors, as this estimation was only intended to compare the outcomes of the first-dose-for-most strategy versus the two-dose strategy, not to determine the priority with which different groups or individuals should receive access to the vaccine. The order of access to the vaccine should therefore be determined independently from the strategy used to distribute vaccinations to those priority groups.

The homogeneity of transmission risk is more limiting, especially under the scenarios where there is a reduction in the transmission of the virus after receiving the vaccine. The ability for certain age groups to be more or less likely to transmit the virus onward was built into the simulator, but data on increased or decreased transmission for different age groups in Canada was not found.

Finally, the effectiveness of vaccination in regards to preventing and transmitting COVID-19 was held constant after an initial period (14 days for the first dose, 7 days for the second). While there is some evidence that the effectiveness remains high throughout the estimated time between doses under both scenarios studied in this paper (up to 13 weeks in the first-dose-for-most scenario), if effectiveness were to drop significantly after a certain point that would have a large effect on the conclusions reported herein.

## METHODOLOGY

Two vaccination strategies are compared in this paper:

Two-dose Strategy

1. Dose 1 given by age-priority in descending 5 year increments
2. Dose 2 given 3 weeks after the initial dose has been administered

First-dose-for-most Strategy

1. Dose 1 given by age-priority in descending 5 year increments

1. Dose 2 given by age-priority in descending 5 year increments after all age groups 15 years and older have received a first dose.

## ASSUMPTIONS

### Vaccination saturation level

For each age group, it’s estimated that a maximum of 95% of the population receives a first or second dose of the vaccine, due either to disqualifying health conditions or vaccine hesitancy. This value was selected based on the coverage goal for childhood vaccinations in Canada^1^.

### Effectiveness period

The effectiveness of one or two doses of the vaccine is assumed to happen in a step-wise fashion, where 14 days after the initial dose or 7 days after the second effectiveness suddenly increases to a new level from 0, for the first dose, or from the first dose effectiveness level, for the second dose. These newer effectiveness levels are then assumed to persist until either a second dose is administered, for the first dose, or permanently, for the second dose.

There are two ways in which this assumption likely over simplifies the effectiveness of the vaccinations. First, it is likely that effectiveness builds over time instead of happening suddenly. Second, there is the potential for a drop in the effectiveness over the course of the study period, in particular for the first dose of the vaccine. The relatively short maximum period between administering the first and second doses of 13 weeks, compared to the interval between many other vaccinations, may negate some of the concern around falling effectiveness.

The step-wise effectiveness assumption described above is used due to the lack of data on the gradient of protection from different doses of the vaccine and in an effort to produce a clear set of results.

### Setting Parameters for the Scenarios

A number of parameters are constant throughout the different scenarios. These include populations in different age ranges, taken from Statistics Canada^2^, the implied case fatality rate, taken from Public Health Ontario^3^, and the maximum number of vaccinations administered per week, based on information released by PHAC. The weekly vaccination numbers begin at 1.5 million doses in week 1, rising to 2 million in week 5, 2.5 million in week 9 and then 3 million in week 13 until the end of the simulation period.

A range of parameters are varied, in particular for the effectiveness of a single dose of vaccine to prevent disease and for the vaccine to prevent transmission of the disease.

For the effectiveness of a single dose of vaccine to prevent disease, the three values compared in this paper are 52.4%^4^, 75%^5^, and 92.6%^6^. These values were selected to cover a range of reported first dose effectiveness based on the available literature.

For the reduction in transmission after vaccination with one or two doses, the values compared are 0%, 50%^7^, 75%^8^, and 89.4%^9^. Again, these values were selected to cover a wide range of possibilities. The values are based on preliminary studies of how effective the Pfizer and AstraZeneca COVID-19 vaccines are at preventing viral transmission, given the limited information available at this time on transmission reduction.

Finally, three parameter values are used for the average number of people a person goes on to infect absent any vaccine effect on transmission. These values are 0.9, 1, and 1.2, covering scenarios where cases are decreasing, steady or increasing.

### Initial Vaccination Rates

Given the April to September period being modelled in the various scenarios, a significant number of vaccinations are presumed to have occurred prior to the start of the scenarios. Based on the vaccine delivery schedule released by PHAC showing 6 million vaccine doses being delivered prior to April first, 95% adults 75 years of age and older are assumed to have previously been fully vaccinated with two doses of vaccine. This corresponds to just under 5.5 million doses of vaccine.

## RESULTS

### General Scenarios

The results from the different scenarios show a range of outcomes where each of the two vaccinations strategies is superior. These results are summarized in the graphic below, with the full results available in the Appendix.

**Figure 1.**
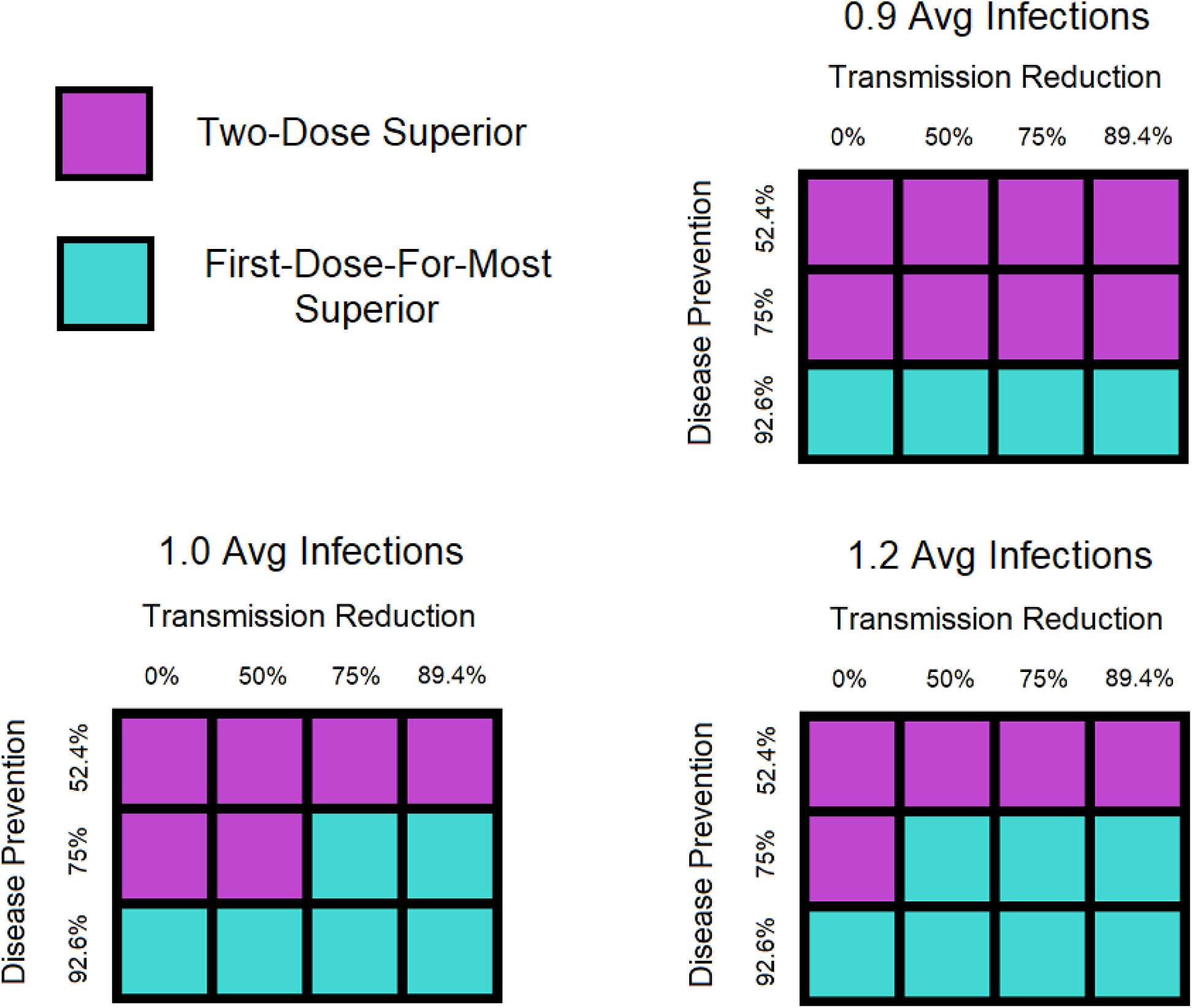
Graphic comparison of results

In general, the results follow an intuitive pattern: when the first-dose effectiveness at preventing disease is set to 92.6%, the first-dose-for-most strategy is superior to the two-dose strategy in every scenario, regardless of whether there is an effect on viral transmission from being vaccinated. Similarly, when the first-dose effectiveness at preventing disease is set to 52.4%, the two-dose strategy is superior to the first-dose-for-most strategy in every scenario.

For the 75% effectiveness value, the results are almost evenly split, depending on the transmission reduction value used and whether cases are increasing, decreasing or steady in the absence of a vaccine-induced effect on viral transmission. In 7 of the 12 scenarios where the 75% disease prevention effectiveness value was used, the two-dose strategy was superior, while in 5 of the 12 scenarios the first-dose-for-most strategy was superior. For this middle value, the two-dose strategy was superior in the decreasing case scenarios and where there was a smaller tested effect for reduction in transmission from vaccination.

One interesting difference between the two vaccination strategies is in the average age at death. The two-dose strategy has a lower average age at death, meaning that proportionately more younger persons would die under the two-dose strategy than using the first-dose-for-most strategy, which could be an important difference if public health officials are looking to maximize life-years saved during the vaccine roll-out.

Additionally, in scenarios where there is any effect on disease transmission from being vaccinated, the first-dose-for-most strategy significantly reduces the number of cases in every scenario. The drop in cases ranges from a low of 7.7% up to a high of 22.6%. This represents a significant reduction in cases, which could also be an important advantage in terms of returning hospital resources to normal usage in preventing and treating diseases other than COVID-19.

### Effect of Strategy Implementation Start Date

To test what effect switching to a first-dose-for-most strategy earlier would have, a scenario was also tested where fewer individuals had received vaccinations prior to the period of study. Using a transmission reduction estimate of 75% and a disease prevention for a first-dose figure of 92.6%, this scenario began with 95% of those over 90 having received two doses of vaccine and 95% of those over 85 having received a single dose of vaccine. This corresponds to just over 1.1 million doses of vaccine administered prior to the study period, approximately the number of vaccine doses administered in Canada by mid-February^10^. Available vaccine doses in each week are assumed to be the same as in earlier scenarios, to allow comparison between the two. Results from this scenario are shown below.

| Two-dose Results |  | First-dose-for-most |  |
| --- | --- | --- | --- |
| Cases | Deaths | Cases | Deaths |
| 438,411 | 1,758 | 371,725 | 1,476 |

In this scenario, the difference in outcomes between the two strategies is even more pronounced. Under the first-dose-for-most strategy, deaths are 16% lower vs. 12.7% lower in the original time-frame and cases are 15.2% lower vs. 11.7% in the original time-frame compared to the two-dose strategy.

### Effect of a Limited Vaccine Supply

Finally, the effect of a reduced vaccine supply on strategy effectiveness was also tested. In this scenario, expected vaccine deliveries per week were halved, a transmission reduction estimate of 75% and a disease prevention for a first-dose figure of 92.6% were used. The starting vaccination rates used were those originally tested, where everyone 75 years and older was assumed to have received two doses of vaccine prior to the study period. Results for this scenario are shown below.

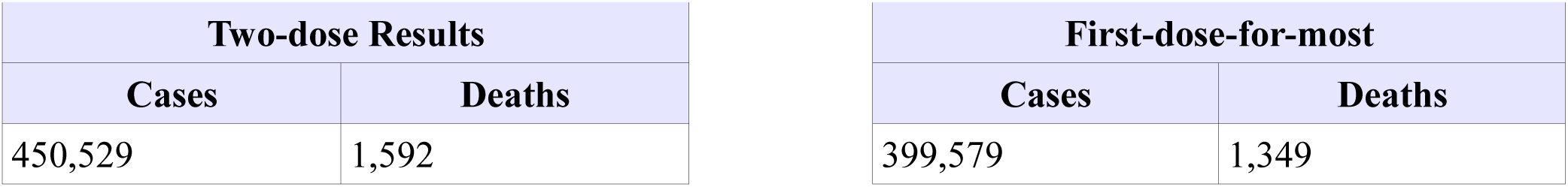

| Two-dose Results |  |
| --- | --- |
| Cases | Deaths |
| 450,529 | 1,592 |

| First-dose-for-most |  |
| --- | --- |
| Cases | Deaths |
| 399,579 | 1,349 |

Once again, the first-dose-for-most strategy is still the superior strategy after reducing the vaccine delivery numbers. In this case, deaths are 15.3% lower and cases 11.3% lower compared to the two-dose strategy.

One important consideration in this scenario is that the maximum gap between vaccinations is significantly longer than under the original scenario at 22 weeks, vs. 13 weeks under the originally tested vaccine delivery schedule. The original assumption that protection from a single dose lasts throughout the study period is more likely to be wrong in this case, and therefore testing that that assumption remained true during the vaccination campaign would be even more important were a reduced-vaccine scenario to occur.

### Additional Considerations

Estimated cases and deaths were for the 24 week period exclusively, and therefore any benefits beyond this period are ignored in this comparison. Under the best-guess scenario with a 75% reduction in transmission of disease, however, the first-dose-for-most strategy cases in the final week were 73.5% lower compared with the two-dose strategy, meaning that cases would likely be significantly higher using the two-dose strategy beyond the period studied.

## Data Availability

All data used in this paper is publicly available.

https://www150.statcan.gc.ca/t1/tbl1/en/tv.action?pid=1710000501

## Appendix - Complete results for the initial set of scenarios.

| Transmission Reduction | Disease Reduction | Total Cases | Total Deaths | Average Death Age |
| --- | --- | --- | --- | --- |
| 0.0% | 52.4% | 470,751 | 1,319 | 65.04 |
| 0.0% | 75.0% | 470,751 | 1,256 | 64.98 |
| 0.0% | 92.6% | 470,751 | 1,212 | 64.92 |
| 50.0% | 52.4% | 307,773 | 1,051 | 64.58 |
| 50.0% | 75.0% | 307,773 | 1,001 | 64.41 |
| 50.0% | 92.6% | 307,773 | 961 | 64.29 |
| 75.0% | 52.4% | 266,020 | 978 | 64.48 |
| 75.0% | 75.0% | 266,020 | 928 | 64.32 |
| 75.0% | 92.6% | 266,020 | 889 | 64.15 |
| 89.4% | 52.4% | 247,675 | 939 | 64.48 |
| 89.4% | 75.0% | 247,675 | 892 | 64.28 |
| 89.4% | 92.6% | 247,675 | 855 | 64.15 |

Double dose – 1.0 average infections w/o vaccine
| Transmission Reduction | Disease Reduction | Total Cases | Total Deaths | Average Death Age |
| --- | --- | --- | --- | --- |
| 0.0% | 52.4% | 840,000 | 1,964 | 64.72 |
| 0.0% | 75.0% | 840,000 | 1,878 | 65.82 |
| 0.0% | 92.6% | 840,000 | 1,812 | 65.87 |
| 50.0% | 52.4% | 452,306 | 1,381 | 64.47 |
| 50.0% | 75.0% | 452,306 | 1,311 | 64.44 |
| 50.0% | 92.6% | 452,306 | 1,258 | 64.34 |
| 75.0% | 52.4% | 368,828 | 1,226 | 64.28 |
| 75.0% | 75.0% | 368,828 | 1,166 | 64.13 |
| 75.0% | 92.6% | 368,828 | 1,117 | 63.98 |
| 89.4% | 52.4% | 334,784 | 1,165 | 64.37 |
| 89.4% | 75.0% | 334,784 | 1,106 | 64.19 |
| 89.4% | 92.6% | 334,784 | 1,062 | 64.05 |

Double dose – 1.2 average infections w/o vaccine
| **Transmission Reduction** | **Disease Reduction** | **Total Cases** | **Total Deaths** | **Average Death Age** |
| --- | --- | --- | --- | --- |
| 0.0% | 52.4% | 3,407,499 | 5,309 | 67.28 |
| 0.0% | 75.0% | 3,407,499 | 5,141 | 67.52 |
| 0.0% | 92.6% | 3,407,499 | 5,012 | 67.69 |
| 50.0% | 52.4% | 1,129,867 | 2,557 | 65.28 |
| 50.0% | 75.0% | 1,129,867 | 2,441 | 65.32 |
| 50.0% | 92.6% | 1,129,867 | 2,347 | 65.37 |
| 75.0% | 52.4% | 783,194 | 2,094 | 64.42 |
| 75.0% | 75.0% | 783,194 | 1,987 | 64.4 |
| 75.0% | 92.6% | 783,194 | 1,904 | 64.35 |
| 89.4% | 52.4% | 661,976 | 1,905 | 64.34 |
| 89.4% | 75.0% | 661,976 | 1,807 | 64.28 |
| 89.4% | 92.6% | 661,976 | 1,730 | 64.15 |

First-dose-for-most – 0.9 average infections w/o vaccine
| **Transmission Reduction** | **Disease Reduction** | **Total Cases** | **Total Deaths** | **Average Death Age** |
| --- | --- | --- | --- | --- |
| 0.0% | 52.4% | 470,751 | 1,626 | 66 |
| 0.0% | 75.0% | 470,751 | 1,344 | 66.02 |
| 0.0% | 92.6% | 470,751 | 1,130 | 65.96 |
| 50.0% | 52.4% | 283,979 | 1,197 | 65.46 |
| 50.0% | 75.0% | 283,979 | 1,019 | 65.13 |
| 50.0% | 92.6% | 283,979 | 878 | 64.77 |
| 75.0% | 52.4% | 244,974 | 1,090 | 65.39 |
| 75.0% | 75.0% | 244,974 | 942 | 64.98 |
| 75.0% | 92.6% | 244,974 | 822 | 64.56 |
| 89.4% | 52.4% | 228,626 | 1,036 | 65.39 |
| 89.4% | 75.0% | 228,626 | 898 | 64.96 |
| 89.4% | 92.6% | 228,626 | 764 | 64.54 |

First-dose-for-most – 1.0 average infections w/o vaccine
| **Transmission Reduction** | **Disease Reduction** | **Total Cases** | **Total Deaths** | **Average Death Age** |
| --- | --- | --- | --- | --- |
| 0.0% | 52.4% | 840,000 | 2,502 | 66.62 |
| 0.0% | 75.0% | 840,000 | 2,044 | 66.83 |
| 0.0% | 92.6% | 840,000 | 1,674 | 67.38 |
| 50.0% | 52.4% | 398,963 | 1,598 | 65.55 |
| 50.0% | 75.0% | 398,963 | 1,323 | 65.31 |
| 50.0% | 92.6% | 398,963 | 1,114 | 64.97 |
| 75.0% | 52.4% | 325,656 | 1,388 | 65.34 |
| 75.0% | 75.0% | 325,656 | 1,165 | 65.03 |
| 75.0% | 92.6% | 325,656 | 997 | 64.59 |
| 89.4% | 52.4% | 297,248 | 1,298 | 65.31 |
| 89.4% | 75.0% | 297,248 | 1,103 | 64.91 |
| 89.4% | 92.6% | 297,248 | 955 | 64.47 |

First-dose-for-most – 1.2 average infections w/o vaccine
| **Transmission Reduction** | **Disease Reduction** | **Total Cases** | **Total Deaths** | **Average Death Age** |
| --- | --- | --- | --- | --- |
| 0.0% | 52.4% | 3,407,499 | 6,937 | 67.61 |
| 0.0% | 75.0% | 3,407,499 | 5,670 | 68.49 |
| 0.0% | 92.6% | 3,407,499 | 4,679 | 69.51 |
| 50.0% | 52.4% | 873,956 | 2,993 | 65.99 |
| 50.0% | 75.0% | 873,956 | 2,372 | 66 |
| 50.0% | 92.6% | 873,956 | 1,876 | 66.08 |
| 75.0% | 52.4% | 609,727 | 2,377 | 65.55 |
| 75.0% | 75.0% | 609,727 | 1,905 | 65.34 |
| 75.0% | 92.6% | 609,727 | 1,539 | 65.05 |
| 89.4% | 52.4% | 524,343 | 2,136 | 65.51 |
| 89.4% | 75.0% | 524,343 | 1,740 | 65.2 |
| 89.4% | 92.6% | 524,343 | 1,429 | 64.83 |

